# The Pediatric Obesity Microbiome and Metabolism Study (POMMS): Methods, Baseline Data, and Early Insights

**DOI:** 10.1101/2020.06.09.20126763

**Authors:** Jessica R. McCann, Nathan A. Bihlmeyer, Kimberly Roche, Cameron Catherine, Jayanth Jawahar, Lydia Coulter Kwee, Noelle E. Younge, Justin Silverman, Olga Ilkayeva, Charles Sarria, Alexandra Zizzi, Janet Wootton, Lisa Poppe, Paul Anderson, Michelle Arlotto, Zhengzheng Wei, Joshua A. Granek, Raphael H. Valdivia, Lawrence A. David, Holly K. Dressman, Christopher B. Newgard, Svati H. Shah, Patrick C. Seed, John F. Rawls, Sarah C. Armstrong

**Author notes:** CONTRIBUTIONS PCS, JFR, SHS, and SCA designed and provided supervision for the study, secured funding, wrote and edited original draft. JRM acted as project manager, curated and analyzed data, wrote and edited original draft. NAB, KR, PCS, JJ, LCK, CBN, JS, and NY curated, analyzed, and interpreted data. SHS, CC, CS, AZ, and JW coordinated clinical activities, collected samples, and curated data. LP, PA, and MA collected samples and curated data. OI, ZW, LAD, and HKD analyzed samples and provided statistical and planning advice. All authors were involved in writing the paper and had final approval of the submitted and published versions.

## Abstract

**Objective:** To establish a biorepository of clinical, metabolomic, and microbiome samples from adolescents with obesity as they undergo lifestyle modification.

**Methods:** We enrolled 223 adolescents aged 10-18 years with Body Mass Index ≥ 95^th^ percentile, along with 71 healthy weight participants. We collected clinical data, fasting serum, and fecal samples at repeated intervals over 6 months. Here we present our study design, data collection methods, and an interim analysis, including targeted serum metabolite measurements and fecal 16S rRNA gene amplicon sequencing among adolescents with obesity (n=27) and healthy weight controls (n=27).

**Results:** Adolescents with obesity have higher serum alanine aminotransferase, C-reactive protein, and glycated hemoglobin, and lower high-density lipoprotein cholesterol when compared with healthy weight controls. Metabolomics revealed differences in branched chain amino acid related metabolites. We also observed differential abundance of specific microbial taxa and lower species diversity among adolescents with obesity when compared with the healthy weight group.

**Conclusions:** The Duke Pediatric Metabolism and Microbiome Study biorepository is available as a shared resource. Early findings suggest evidence of a metabolic signature of obesity unique to adolescents, along with confirmation of previously reported findings describing metabolic and microbiome markers of obesity.

**Clinical Trial Registration:** Biorepository: NCT02959034

Observational Trial: NCT03139877

**What is already known about this subject?:**

- The intestinal microbiome plays an important role in adult obesity and regulation of metabolism. Although it is well-established that obesity has its roots in childhood, very little is known about the role of the microbiome in pediatric obesity and how it changes during adolescence.

**What are the new findings in your manuscript?:**

- This manuscript provides details of a new shared biorepository including clinical data, stool samples and plasma samples from a diverse cohort of 223 adolescents with obesity followed longitudinally over 6 months during a weight management intervention, as well as 71 adolescents with healthy weight as a comparison group.
- Interim analyses suggest that adolescents with obesity have microbiome signatures and metabolite profiles similar to adults, however key differences in microbial communities and metabolic by-products are identified.

**How might your results change the direction of research or focus of clinical practice?:**

- The POMMS biorepository will be available for investigators to use in future research, to elucidate the underlying mechanisms of obesity and related chronic health conditions.
- Preliminary data reveal metabolite profiles that suggest adolescence may be a window of metabolic plasticity and disease reversibility
- Microbiome and metabolomic signatures suggest potential biomarkers that may serve as prognostic or predictive factors in disease remission, or targets for future therapeutics.

## INTRODUCTION

Despite sustained research into effective treatment strategies, pediatric obesity remains at epidemic levels, and strongly predicts adult obesity, metabolic, and cardiovascular disease (1). Currently, one in three children in the US are classified as overweight or obese, with the highest prevalence among low-income and non-white youth (2). The US Preventative Service Task Force recommends that pediatric providers screen all children aged 6-18 years for obesity annually, using the Centers for Disease Control and Prevention sex- and age-specific Body Mass Index (BMI) curves. Additionally, children with BMI at or above the 95^th^ percentile should be referred to a comprehensive behavioral intervention of medium to high intensity, defined as achieving ≥26 hours of contact over six months (3). However, existing data show that recommended interventions for pediatric obesity result in a heterogeneous response, with most participants showing non-significant BMI reduction (4), with little understanding of the underlying predictive factors for treatment success.

Strong evidence suggests that the intestinal microbiome and its products influence chronic diseases such as obesity, insulin resistance, and heart disease (5). Observational studies in adult humans and stool transplantation experiments in animal models have found novel connections among obesity, insulin resistance, metabolic pathways, inflammation, and intestinal microbiota that may differ between individuals (6). Circulating and tissue metabolites serve as diagnostic, prognostic, and therapeutic targets for metabolic disease, and can be affected by microbiome composition (7). Thus, analyses of the microbiome and metabolome have the potential to provide insights into the host-microbiome pathways and inform personalized and effective treatments.

Adolescents provide a unique opportunity to garner deeper insights into the obesity-associated microbiome and related metabolic pathways during the transition from childhood to adulthood. First, adolescents with severe obesity mimic adult metabolic and cardiovascular risk phenotypes (8, 9). However, unlike adults, adolescents are at early stages of disease, have fewer and less severe co-morbid conditions, and tend to be treatment-naïve. Second, adolescence is a unique window of development and its associated microbiome and metabolic signatures in health and disease are understudied compared to those signatures in adults (10). Similarly, while significant evidence supports the association between decreased gut microbiome diversity and obesity in adults, this has not been confirmed in a larger and racially diverse group of adolescents (11). Third, the prospective longitudinal design of adolescents enrolled in weight mangagment will provide insights into metabolomic and microbiome changes in response to treatment and the early biomarkers of improved health. Last, while obesity and its complications are more prevalent in racial and ethnic minorities, most studies to date have not enrolled significant numbers of these groups, limiting our ability to treat the most at-risk populations (12).

The Pediatric Obesity Microbiome and Metabolism Study (POMMS) is an NIH-funded prospective observational study with two main objectives: 1) to identify serum metabolic biomarkers or stool microbial signatures in adolescents for obesity severity, obesity-related comorbid disease, and the later development of obesity-related disease; and 2) to identify serum or stool biomarkers that predict response to treatment, precede response to treatment, or that may help to personalize treatment to optimize individual outcomes. We describe herein the POMMS methods and procedures, as well as early findings from an interim analysis of 27 participants with obesity and 27 healthy weight controls.

## METHODS

### Screening and enrollment

Research coordinators screened patients referred by their primary care physician to the Duke Healthy Lifestyles clinic in Durham, NC for obesity treatment. Study inclusion criteria were age 10-18 years, Sexual Maturity Rating ≥ 2, age- and gender-specific BMI ≥ 95^th^ percentile, ability to communicate in English or Spanish, and plans to stay in weight management for ≥ 6 months. Patients were excluded for recent weight loss of ≥ 5%, antibiotic use in the prior month or anticipated in the next 6 months, immunodeficiency, prior transplantation, Type 1 diabetes mellitus, inborn errors of metabolism, endogenous obesity (i.e. hyperthyroidism, Cushing syndrome, Mc4R mutation), drug-induced obesity (steroids or anti-psychotics), a significant medical or mental health condition, or pregnancy. Healthy weight controls were recruited from the same community health system, aged 10-18 years with a Sexual Maturity Rating ≥ 2, and age- and gender-specific BMI ≥5^th^ and <85^th^ percentile.

### Study design

The study design and procedures were approved by the Duke Institutional Review Board. Eligible participants had parental consent and child assent for the observational study and long-term storage and future use of clinical data and biospecimens. Figure 1 highlights the prospective observational treatment group assignment and study assessments.

**Figure 1.**
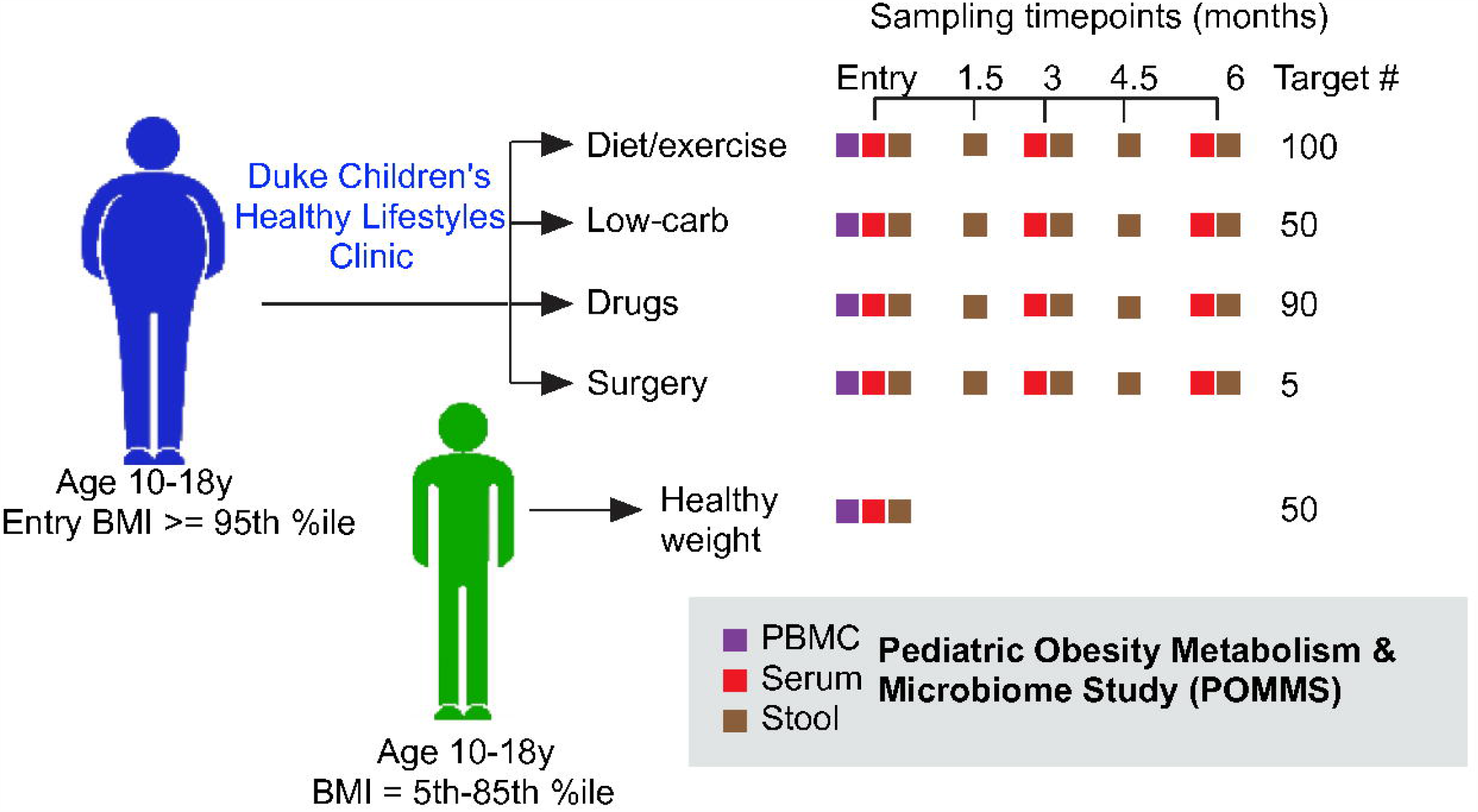
POMMS study design. Adolescents between 10 and 18 with either BMI >5^th^ %ile to < 85^th^ percentile (healthy weight controls, or HWC) or BMI equal to or over the 95^th^ percentile (adolescents with obesity, or OB) were recruited to join the study. Target numbers of recruits for each intervention are indicated, along with actual recruitment numbers. Age matched HWC recruits had clinical measurements of health as well as serum, stool, and PBMCs collected at a single timepoint, while OB recruits were evaluated and offered a treatment arm that was clinically relevant depending on the individual. OB recruits then had clinical measurements of health, blood, PBMC and stool samples taken at baseline, 3 months, and 6 months (or termination) of the study. Stool samples were also collected from OB patients at 1.5 and 4.5 months after the first visit. All samples are stored in replicates in the POMMS Biorepository managed within the Duke Children’s Biobank to be made available for research purposes.

### Groups

All participants in the <u>intervention (obese or OB) group</u> are patients in the Healthy Lifestyles program, which includes visits to a multi-disciplinary clinic and membership in a community-based fitness program (described below and in Supplemental Methods). All participants in the <u>comparison (healthy weight or HWC) group</u> received a single measurement at baseline only and no additional intervention.

<u>The Healthy Lifestyles clinic</u> is a tertiary-care weight management program that meets current recommendations for family-based child and adolescent obesity treatment (13). The Healthy Lifestyles clinic sees 800 new patients aged 2-18 annually, and more than 15,000 children have received care to date with demonstrated efficacy (14). Advanced treatment options include a low-carbohydrate diet (<30g carbohydrate/day), weight-loss medications (e.g. metformin, phentermine, lisdexamphetamine, lorcaserin, and topriamate), and surgery (Roux-en-Y Gastric Bypass or Laparascopic Vertical Sleeve Gastrectomy). Shared decision-making between parent, teen, and healthcare provider guides treatment choice, and decisions are made based on child’s age, obesity severity, comorbidities, and family preference. In addition to clinical care, participants are enrolled in a <u>community-based fitness program</u>, delivered through a local Parks and Recreation Department (see Supplemental Methods for details).

### Measures

<u>The primary study outcome</u> was change in BMI at 3 and 6 months and was determined through standardized measures of body weight and height measured at baseline, 3 and 6 months using a digital scale and stadiometer. A large number of children referred to the Healthy Lifestyles clinic have a BMI significantly greater than the 95^th^ percentile. Therefore, as recommended by Flegal *et al*. (15) we assess and report raw BMI, BMI percentile, and child relative BMI, expressed as a standard deviation score (zBMI) and as a percent of the BMI value at the 95^th^ percentile (%95^th^ BMI) to evaluate and track obesity.

<u>Secondary endpoints</u> were collected at baseline, 3 and 6 months. Cardiorespiratory fitness was assessed by heart rate at the completion of the YMCA submaximal bench-stepping test (16). Blood pressure was measured with a calibrated auscultatory sphygmomanometer in the seated position with an appropriately sized cuff using standard methods (17). Fasting blood samples were obtained for measuring cardiometabolic biomarkers, including lipids, glucose, and transaminases as described in Supplemental Methods. Body fat percentage was measured using calibrated bioelectrical impedance. Parent BMI was directly measured from parent height and weight using calibrated stadiometers.

<u>Survey data</u> (available in English and Spanish) included weight-specific quality of life, measured using the validated “Sizing Me Up” child obesity-specific instrument at baseline and 6 months (18). Household income, transportation access, parent stress, food insecurity, home food environment, and parental expectations for treatment were also collected at baseline (Supplemental Methods). Factors likely to influence the intestinal microbiome, including birthweight, breastfeeding history, pets in the home, recent or distant antibiotic use, concurrent medication use, and other self-reported measures were recorded.

#### Serum

Following a fast of 8-14 hours, blood was collected in anti-coagulant containing tiger top (Becton Dickenson (BD) #367988) and green top (BD #367874) tubes at baseline, 3 and 6 months. Blood samples were processed within 2 hours of collection according to established protocols (19). Serum samples were then snap frozen in liquid nitrogen and stored at −80°C in the Duke Children’s Biobank.

#### Stool

Patients collected stool samples and immediately stored the collection container (Fisher Scientific #02-544-208) in their home freezer. Patients then returned the sample or scheduled a home pickup within 18 hours of stooling. Stool was transported frozen and transferred to a −80°C freezer for long-term storage. Stool samples for analysis were processed by thawing on ice in a biological safety cabinet, and opened to the atmosphere for a maximum of 10 minutes while samples were portioned into approximately 2g aliquots. Aliquots were then stored at −80°C in the Duke Children’s Biobank.

Serum and stool samples were organized into a POMMS Biorepository that is available for researchers, with complete clinical data, and fecal and serum longitudinal sample sets from 50 HWC and 140 OB and baseline data and samples from over 200 adolescents with obesity. More information on the repository, sample requests, protocols, recruitment and study completion incentivizing strategies can be found at https://sites.duke.edu/pomms/.

### Interim analysis

An interim group of baseline samples in the POMMS Biorepository was selected for an initial analysis of association between metabolites and microbiome characteristics and adolescent obesity. These interim analysis samples were taken at the study enrollment visit from early study entrants with complete sample sets and included 27 healthy weight controls (HWC) and 27 adolescents with obesity (OB) matched for puberty status, sex, and ethnicity.

### Targeted serum metabolite analysis

#### Sample preparation and data acquisition

74 amino acids, acylcarnitines and organic acids were analyzed by targeted mass spectrometry methods employing stable isotope dilution for analyte quantitation. Amino acids and acylcarnitine were measured by flow injection tandem mass spectrometry using sample preparation methods described previously (20). The data were acquired using a Waters TQD mass spectrometer equipped with AcquityTM UPLC system and controlled by MassLynx 4.1 operating system (Waters, Milford, MA). Organic acids were quantified as described (21), employing Trace Ultra GC coupled to ISQ MS operating under Xcalibur 2.2 (Thermo Fisher Scientific, Austin, TX).

#### Statistical analysis

Levels of all targeted metabolites were log-transformed and tested for association with obesity using a linear model, adjusting for baseline BMI, age, sex, race, and ethnicity and correcting for false discovery rate (22). The complete set of metabolite data can be found online (Supplemental Data Table 1).

**Table 1.**
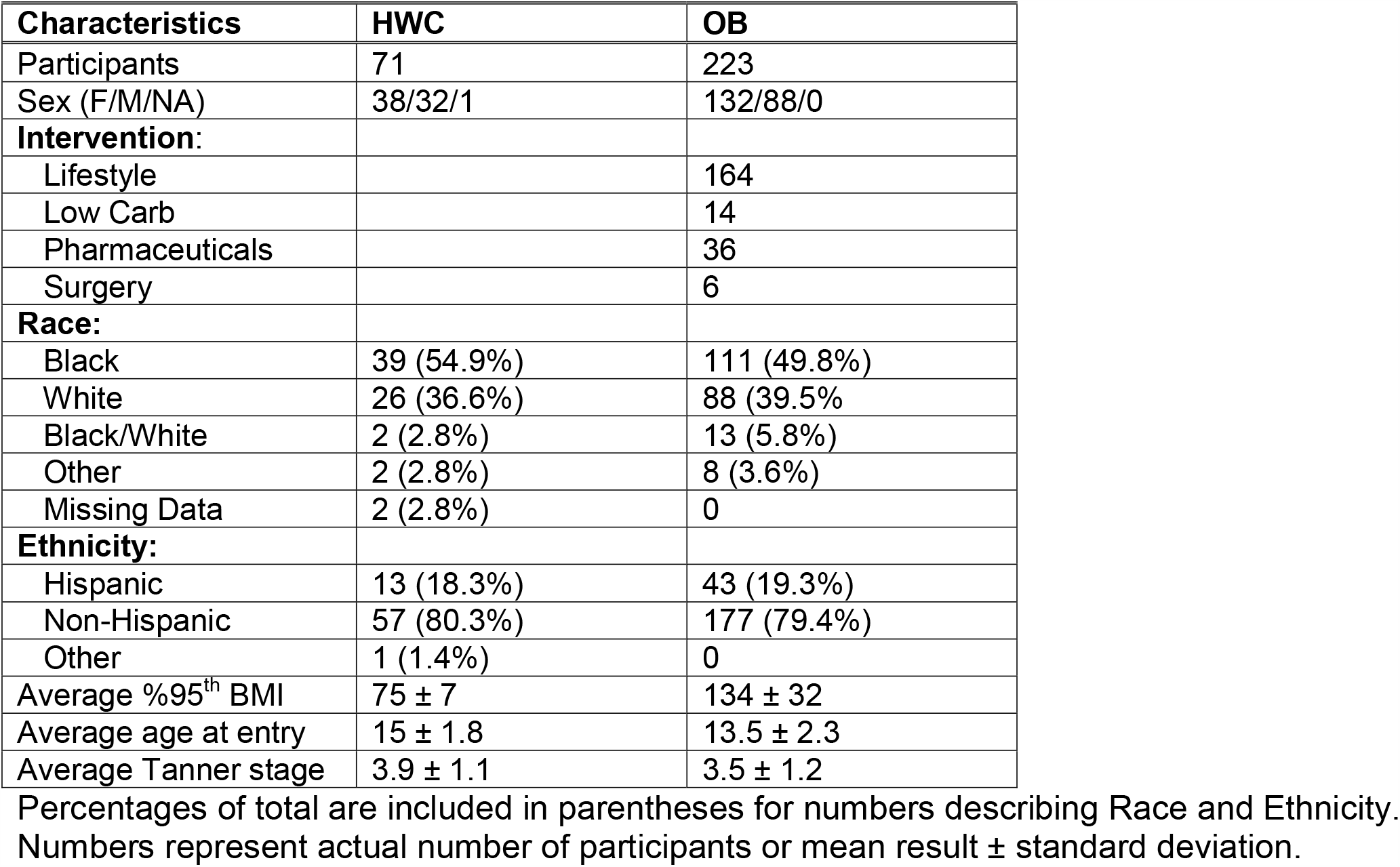
Description of all study participants in the Healthy Weight Control (HWC) and Adolescents with Obesity (OB) cohort.

| Characteristics | HWC | OB |
| --- | --- | --- |
| Participants | 71 | 223 |
| Sex (F/M/NA) | 38/32/1 | 132/88/0 |
| **Intervention:** |  |  |
| Lifestyle |  | 164 |
| Low Carb |  | 14 |
| Pharmaceuticals |  | 36 |
| Surgery |  | 6 |
| **Race:** |  |  |
| Black | 39 (54.9%) | 111 (49.8%) |
| White | 26 (36.6%) | 88 (39.5%) |
| Black/White | 2 (2.8%) | 13 (5.8%) |
| Other | 2 (2.8%) | 8 (3.6%) |
| Missing Data | 2 (2.8%) | 0 |
| **Ethnicity:** |  |  |
| Hispanic | 13 (18.3%) | 43 (19.3%) |
| Non-Hispanic | 57 (80.3%) | 177 (79.4%) |
| Other | 1 (1.4%) | 0 |
| Average %95<sup>th</sup> BMI | 75 ± 7 | 134 ± 32 |
| Average age at entry | 15 ± 1.8 | 13.5 ± 2.3 |
| Average Tanner stage | 3.9 ± 1.1 | 3.5 ± 1.2 |

Percentages of total are included in parentheses for numbers describing Race and Ethnicity. Numbers represent actual number of participants or mean result ± standard deviation.

### 16S rRNA gene sequencing

#### DNA purification, amplification, and sequencing

The Duke Microbiome Shared Resource staff extracted bacterial DNA from stool sample aliquots (∼120 mg stool per sample) using a MagAttract PowerSoil DNA EP Kit (Qiagen, 27100-4-EP) and a Retsch MM400 plate shaker. Resulting DNA concentration was assessed using a Qubit dsDNA HS assay kit (ThermoFisher, Q32854). Bacterial community composition in isolated DNA samples was characterized by amplification of the V4 variable region of the 16S rRNA gene by PCR using the barcoded primers 515 forward and 806 reverse following the Earth Microbiome Project protocol (http://www.earthmicrobiome.org/). Equimolar 16S rRNA PCR products from all samples were pooled prior to sequencing by the Duke Sequencing and Genomic Technologies shared resource on an Illumina MiSeq instrument configured for 250 base-pair paired-end sequencing runs. Raw sequence data can be found at the NIH Sequence Read Archive (submission in progress). Resulting data were processed using the Qiime2 bioinformatics platform (23). Data denoising was conducted with the DADA2 (24), Bioconductor (25) package in the R statistical programming environment. The RAxML algorithm (26) was used to generate the phylogenetic tree using a midpoint-rooting methodology. We used the Silva 132 classifier for taxonomic classification using variable region 4 (99% identity) (27).

#### Statistical analysis

Statistical analysis of taxonomic data, including relative abundance plots, alpha and beta diversity comparisons, and PCoA plots, were performed using the R packages Phyloseq v1.26.1, vegan 2.5.5 (28, 29), Phylogenetic Isometric Log-Ratio (PhILR) transformation (30), and LefSe (31). Relevant R code can be found at github (submission in progress).

## RESULTS

We enrolled 294 adolescents (223 adolescents with obesity, or OB, and 71 healthy weight control adolescents, or HWC) (Table 1) between December, 2016 and July, 2019. Approximately half (49.8% OB v. 54.9% HWC) of all participants identified as Black, while 19.3% of the OB group and 18.3% of the HWC group identified as Hispanic. Demographic characteristics of the interim analysis sample (27 OB and 27 HWC study participants) are shown in Table 2. Traditional laboratory parameters, including triglycerides, insulin, total cholesterol, blood pressure, and high- and low-density lipoprotein cholesterol (HDL and LDL, respectively) were analyzed and compared between the healthy weight cohort (HWC) and adolescents with obesity (OB) (All parameters collected are described in Supplemental Methods). Triglycerides, fasting glucose, LDL, and total cholesterol were not significantly different between HWC and OB after adusting for false discovery rates (FDR) (Table 2). We did find higher levels of homeostatic model of insulin resistance (HOMA-IR (32), HDL, glycated hemoglobin (Hb1Ac), C-reactive protein (CRP, Fig. 2), and alanine amino transferase (ALT, Fig. 2) in the OB as compared to the HWC control group, and lower levels of HDL cholesterol (Table 2).

**Table 2.**
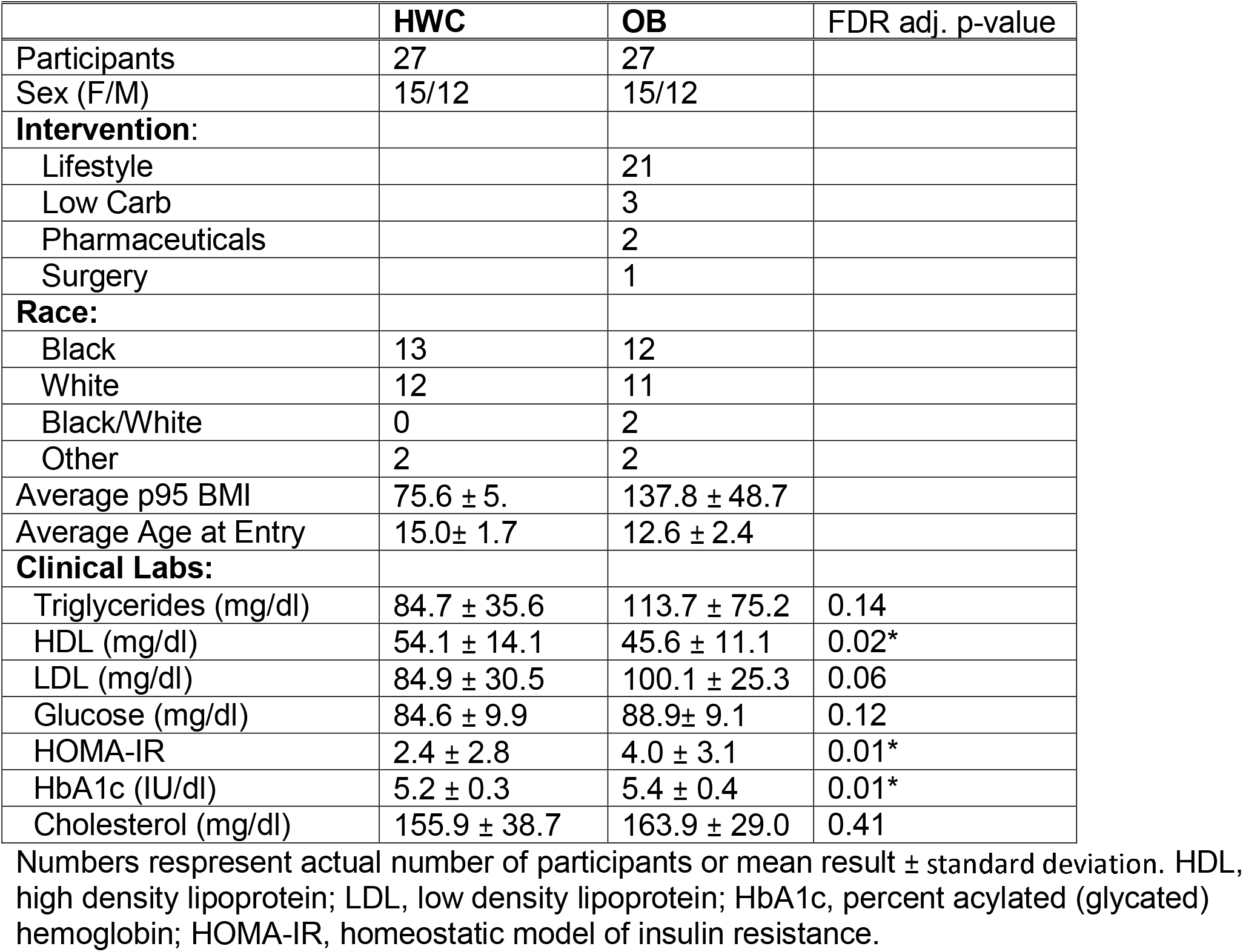
Demographics and clinical lab results for study participants included in the interim analysis.

|  | **HWC** | **OB** | FDR adj. p-value |
| --- | --- | --- | --- |
| Participants | 27 | 27 |  |
| Sex (F/M) | 15/12 | 15/12 |  |
| **Intervention:** |  |  |  |
| Lifestyle |  | 21 |  |
| Low Carb |  | 3 |  |
| Pharmaceuticals |  | 2 |  |
| Surgery |  | 1 |  |
| **Race:** |  |  |  |
| Black | 13 | 12 |  |
| White | 12 | 11 |  |
| Black/White | 0 | 2 |  |
| Other | 2 | 2 |  |
| Average p95 BMI | 75.6 ± 5. | 137.8 ± 48.7 |  |
| Average Age at Entry | 15.0± 1.7 | 12.6 ± 2.4 |  |
| **Clinical Labs:** |  |  |  |
| Triglycerides (mg/dl) | 84.7 ± 35.6 | 113.7 ± 75.2 | 0.14 |
| HDL (mg/dl) | 54.1 ± 14.1 | 45.6 ± 11.1 | 0.02* |
| LDL (mg/dl) | 84.9 ± 30.5 | 100.1 ± 25.3 | 0.06 |
| Glucose (mg/dl) | 84.6 ± 9.9 | 88.9± 9.1 | 0.12 |
| HOMA-IR | 2.4 ± 2.8 | 4.0 ± 3.1 | 0.01* |
| HbA1c (IU/dl) | 5.2 ± 0.3 | 5.4 ± 0.4 | 0.01* |
| Cholesterol (mg/dl) | 155.9 ± 38.7 | 163.9 ± 29.0 | 0.41 |

Numbers represent actual number of participants or mean result ± standard deviation. HDL, high density lipoprotein; LDL, low density lipoprotein; HbA1c, percent acylated (glycated) hemoglobin; HOMA-IR, homeostatic model of insulin resistance.

**Figure 2.**
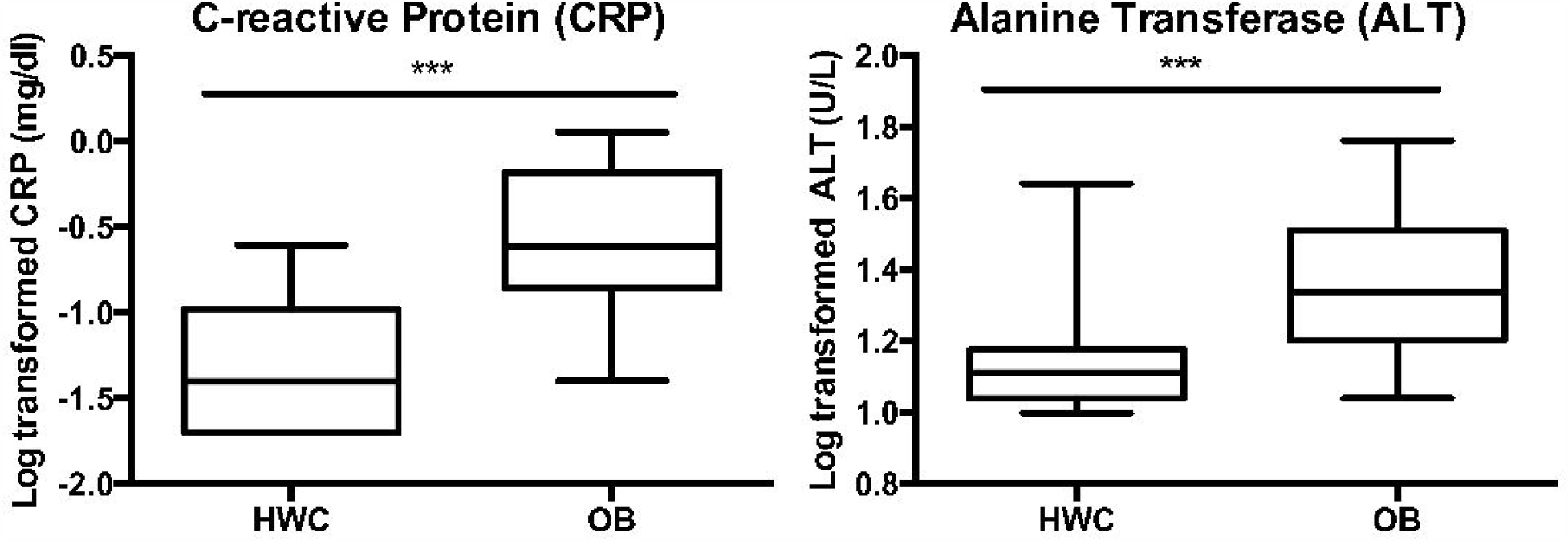
Clinical lab measurements of liver function and inflammation were significantly higher in OB v HWC. Log transformed serum values of alanine transferase (ALT) and C-reactive protein (CRP) were compared at baseline between healthy weight control (HWC) and adolescents with obesity (OB). Error bars indicate standard deviation of the mean.*** = P< 0.001 following correction for multiple testing. See Methods for details on statistical analysis.

### Metabolic Profiling

After FDR adjustment for multiple comparisons, no metabolites were significantly different between the OB and HWC groups. However, several metabolites related to branched chain amino acid (BCAA) metabolism were nominally significantly different (Fig 3). Specifically, the BCAA valine (Fig. 3A) was higher in OB vs. HWC, while the ketoacid products of BCAA catabolism, α-ketoisocaproate (KIC) and α-keto-β-methylvalerate (KMV), were lower in OB vs HWC (Fig. 3B-C). Serum glycine (Fig. 3D) was also lower in OB when compared with HWC. We also observed higher serum glycerol and serum insulin in the OB vs. HWC group (Fig 3E-F).

**Figure 3.**
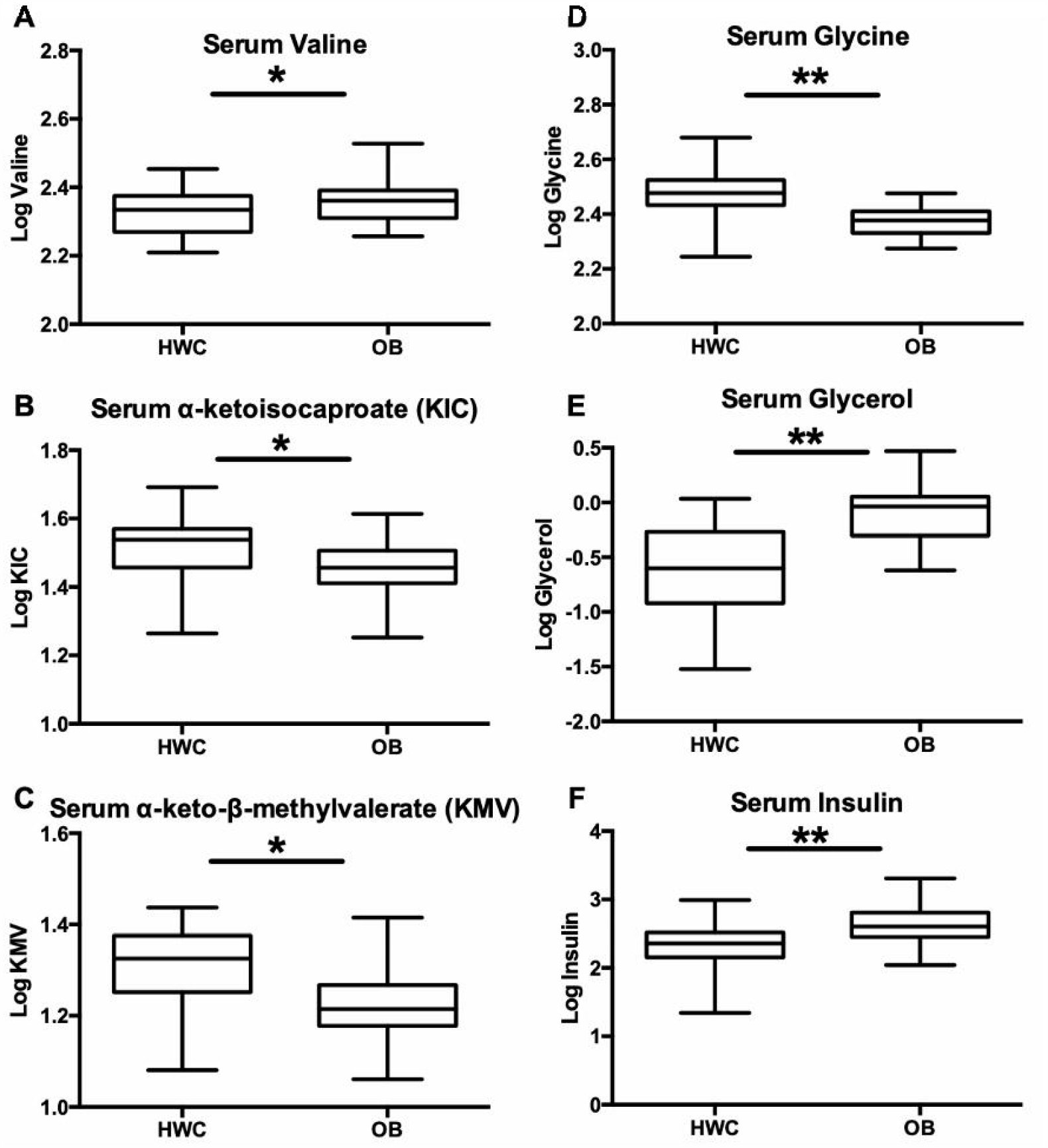
Serum metabolites associated with obesity in adults were significantly different in our adolescent interim cohort at baseline. Log transformed serum metabolite values are presented. Error bars indicate standard deviation of the mean. *, p<0.05. **, p<0.01. See Methods for details on statistical analysis.

### Microbiome Profiling

Stool sample 16S rRNA gene sequence analysis revealed significant differences in measurements of alpha and beta diversity between OB and HWC groups (Fig. 4A, and Supplemental Fig. S1). However, we did not detect significant differences in alpha or beta diversity based on sex nor on age at sampling, suggesting that obesity as defined by %95^th^ BMI was the main driver of the observed difference in microbiome diversity between groups (data not shown).

**Figure 4.**
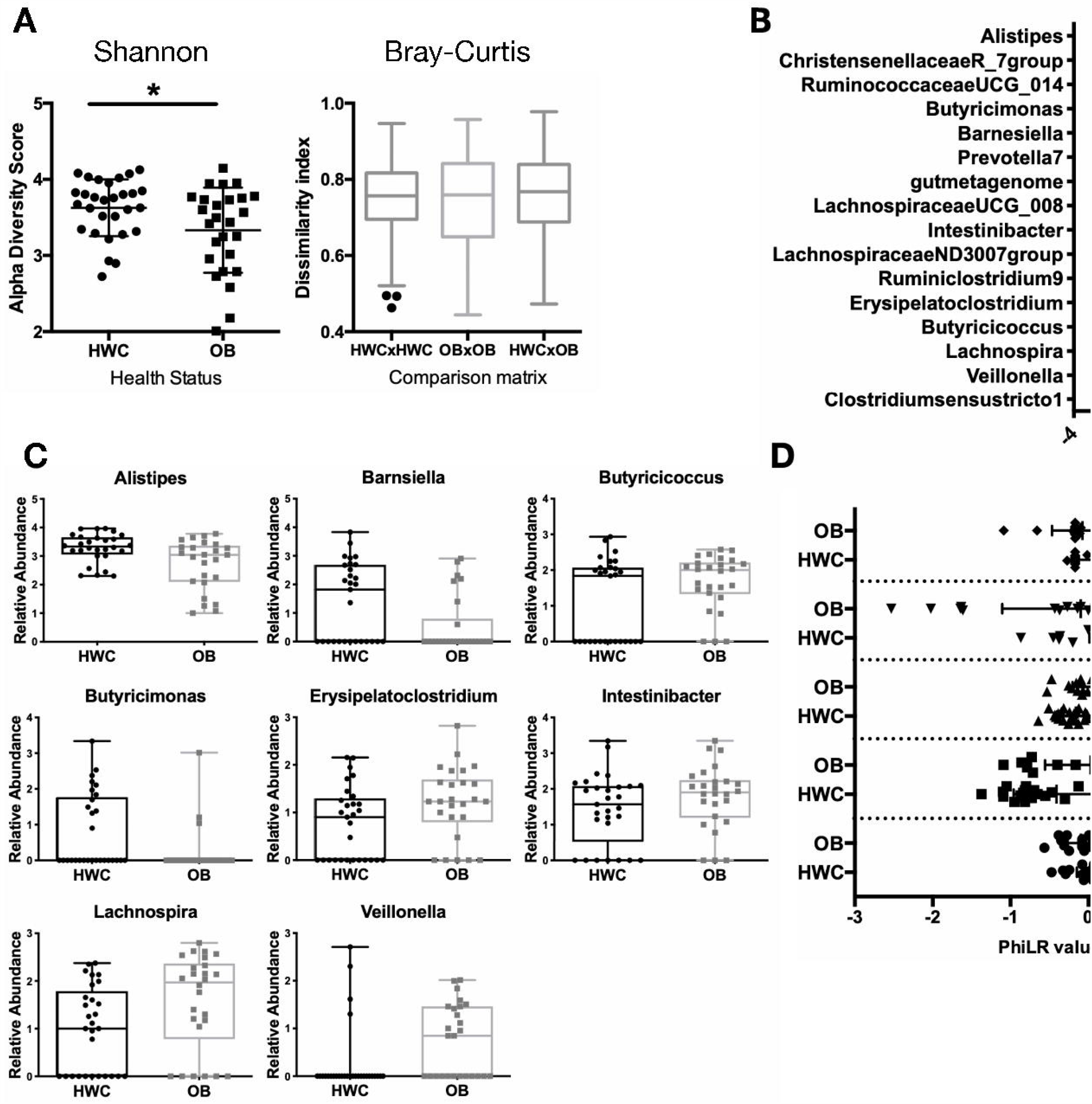
A subset of adolescents with obesity have reduced gut microbial diversity and differences in abundance of specific microbial taxa when compared to their healthy weight counterparts. **(A)** Shannon α-diversity and Bray Curtis β-diversity metrics between HWC and OB stool 16S rRNA gene sequences based on identified amplicon sequence variants. (**B-C**) The Linear discriminant analysis (LDA) effect size tool (LEfSe) was applied to taxonomic data resulting from 16S rRNA gene amplicon sequencing. (**B**) Relative abundance of taxa with that vary significantly by cohort are displayed. (**C**) Count data of each taxon with a significantly different abundance between HWC and OB cohorts in the LDA analysis. Each symbol represents abundance in an individual sample. **(D**) PhiLR Taxa identified by 16S rRNA gene sequencing that were present at a read count of > 3 across at least 10% of patient samples were retained for analysis. PhiLR results are displayed as log relative abundances of opposing clades in a phylogenetic tree and significant balances were chosen on the basis of a simple predictive model. A PhILR value greater than zero indicates greater abundance of the numerator component relative to the denominator component and a negative value indicates the reverse. These balances were most effective at predicting patient membership in either the HWC or OB cohort. See Methods section for details on statistical analyses and access to relevant R code.

There were several bacterial taxa (here defined as a group of one or more populations of an organism or organisms that forms the lowest phylogenenetic unit identifiable by 16S rRNA gene sequencing) detected as differentially abundant in OB vs. HWC groups following analysis using the linear discriminant analysis (LDA) effect size tool (LEfSe) (Fig. 4C-D). Members of the Christensenellaceae and Ruminococcae UCG_14 families, along with an *Alistipes* species, were more likely to be found in samples from HWC, while two Lachnospiraceae families and a *Lachnospira* species were more likely to be associated with OB samples. To identify microbiome community configurations that might be associated with obesity, we examined the phylogenetic relatedness of microbes in each group using 2 different methods. In the first, we applied the Phylogenetic Isometric Log-Ratio (PhILR) transformation (Fig. 4D) to select taxonomic balances which were most effective at predicting patient membership in either the HWC or OB group. The abundance of *Eubacterium brachy* genera relative to the abundance of the Family XIII AD3011 group was higher in the HWC group as compared with OB. Conversely, the abundance ratio of Lachnospiraceae family members to *Ruminococcus gnavus* was substantially lower in the OB group as compared to HWC (Fig. 4D).

In the second phylogenetic analysis, species or lowest taxa relatedness was determined without regard to weight status (Fig. 5A). The resulting five phylogenetic clusters were then applied to weight status, revealing groups of related organisms that were then color coded and plotted. We found that some clusters were more likely to appear in the HWC cohort than the OB cohort, and vice versa (Fig. 5B). Taxa associated with the red cluster were more likely to be found in the OB group when compared with HWC, and this cluster appears to be driven by the decreased abundance of *Bacteroides* family members. Increased abundance of several Ruminococcae and specific members of the Provetellaceae was also more likely to be associated with OB (Fig. 5B and Supplemental Fig. S2).

**Figure 5:**
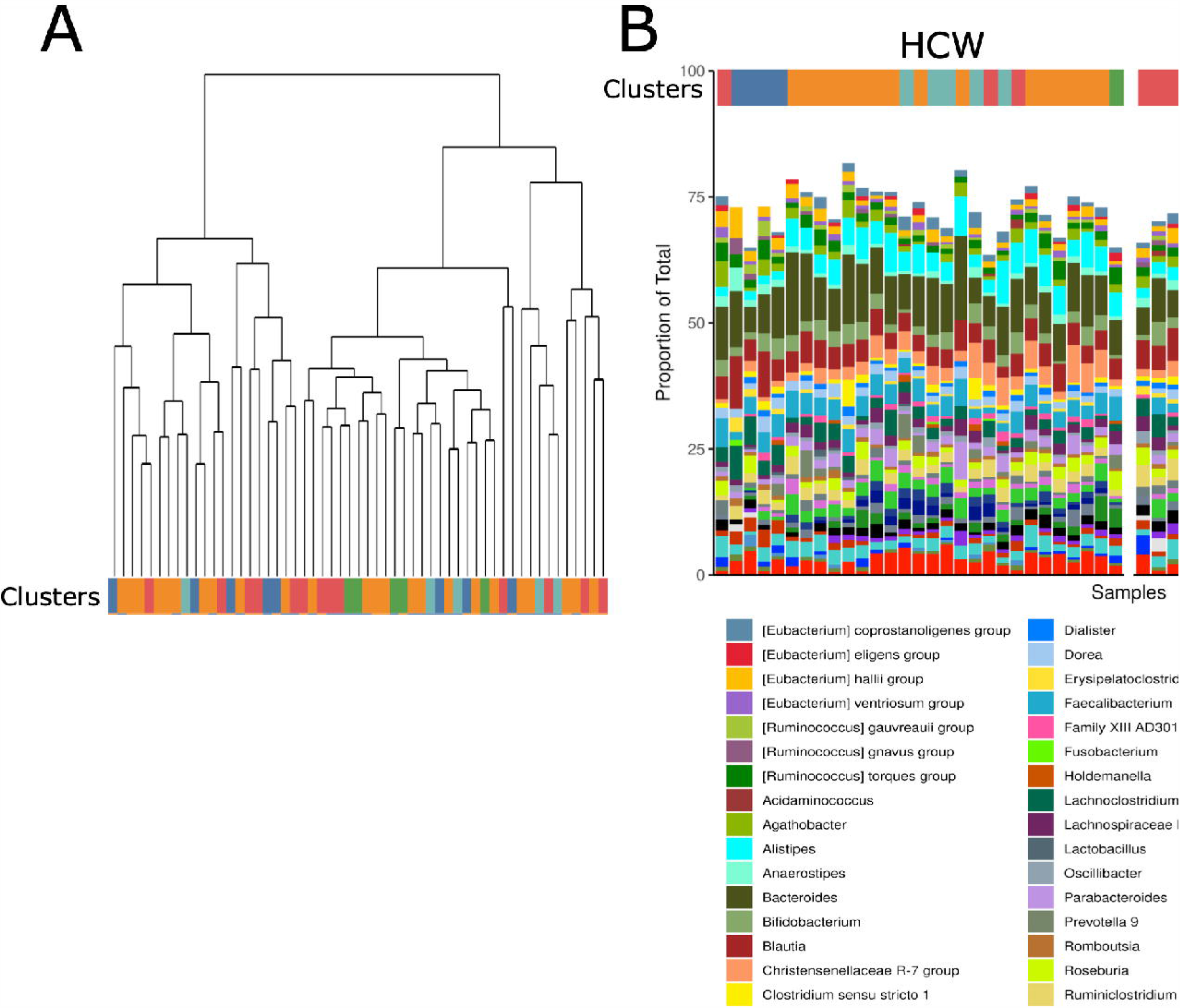
Phylogenetic clustering of gut microbial taxa suggest that genetic relationships predict presence or absence of taxa in OB cohorts. **(A)** Taxa identified by 16S rRNA gene amplicon sequencing were subjected to unsupervised clustering into 5 taxonomic clusters and grouped by color and treatment status. Each terminal node of the tree represents results from an individual fecal sample. **(B)** Clusters were assigned a color and each fecal sample result was grouped by cluster in (A) and then divided by HWC or OB status in order of participant ID number. See methods for links to relevant R code and raw data.

## DISCUSSION

In this interim analysis, we demonstrated proof of concept for 1) collection of clinical data, plasma and stool samples from a racially and ethnically diverse sample of adolescents with and without obesity, and 2) for conducting the analyses that may help to untangle the associations between microbiome and metabolome that contribute to health and disease. While representative of a small subset of the POMMS cohort, we observed interesting findings that both support and contradict existing adult data, and deserve further investigation.

### Clinial lab measures are inconsistently effective at distinguishing obese vs. non-obese

We did not detect significant differences in some laboratory screening measures for obesity co-morbidities between participants in the obese and non-obese cohorts. This is not surprising, in part because of the small sample size, but also because larger studies have demonstrated that comorbidities in children present most clearly at the extremes of excess weight (33). Future analysis of the full POMMS cohort will be appropriately powered to confirm or refute this finding, however this early result suggests that some clinical labs may not be the most sensitive markers of early cardiovascular disease or diabetes. In contrast, levels of the inflammatory marker CRP and the liver function enzyme ALT were significantly higher in adolescents with obesity when compared to the healthy weight cohort. In a recent meta-analysis of pediatric weight loss trials, reduction in CRP and ALT scores were significantly correlated with BMI reduction over time (34). However, the fact that our interim OB participants are younger than the HWC due to pubertal stage at sampling (Table 2) should be considered when evaluating this data.

### Targeted serum metabolite analysis confirms BCAA biomarkers of adult obesity are also present in adolescence, but demonstrates potential unique metabolic plasticity

Analysis of targeted serum metabolite levels between HWC and OB groups revealed additional interesting results. We found that the branched-chain amino acid (BCAA) valine and the related branched chain ketoacids (BCKA) KMV and KIC were found at differing levels between OB and HWC cohorts, but only valine was positively associated with OB, while the BCKA were negatively associated with OB (Fig. 4). In addition, glycine levels were lower in our OB adolescents, consistent with studies in obese adults and rodent models (35) (Fig. 3D). These metabolites were analyzed in our adolescents as they are emerging biomarkers for obesity and future development of type 2 diabetes in adults (10). In fact, BCAA are typically elevated in adults with obesity and are among the strongest baseline predictors of improvements in insulin sensitivity with weight loss (36) as well as later life development of type 2 diabetes in adults (10). However, unlike in adult obesity in humans and animals models, where BCAA and BCKA are both elevated, BCKA were lower in OB subjects from our adolescent cohort. This may be further evidence of an existing hypothesis that there is “metabolic plasticity” in youth protecting against insulin resistance by preventing mitochondrial overload of BCAA metabolites (10). Low plasma glycine levels may also be predictive of type 2 diabetes development, and plasma glycine levels increase with an improvement in insulin sensitivity (37).

Evidence suggests that levels of some metabolites such as circulating BCAA and fatty acid oxidation intermediates correlate with insulin resistance and higher HOMA-IR scores can be influenced by sex (38). To underscore this point, in a recent metabolomic survey of overweight and healthy weight children, BCAA and related metabolites trended as significantly different in obesity until the data was corrected for age, sex, and sexual maturity stage (39). However, in other larger studies, serum or plasma BCAA were significantly positively associated with obesity over a large age range of children and young adults (10). Indeed, in these larger studies with a combined sample size of over 1,000 HWC and OB children, the principal components with the largest contribution to risk factors of later cardiometabolic disease such as HOMA-IR and BMI were composed of the BCAA and their breakdown products. Thus, our data agree with these prior studies in that that elevated BCAA levels were associated with obesity in adolescents, and may provide rationale for more research into their viability as biomarkers of later life disease development (40). A recent attempt to identify such biomarkers of later life disease progression associated with obesity in 396 girls (mean age, 11.2 years at baseline) over a 7 year period demonstrated that serum levels of the BCAA leucine and isoleucine at baseline positively correlated with triglyceride levels at age 18, regardless of the triglyceride level at baseline (10).

Elevated glycerol levels in our OB cohort (Fig. 3E) suggest that adolescents with OB have increased levels of lipolysis. Lipolysis is elevated in adult obesity, resulting in higher circulating glycerol levels in adults, and in a recent study, partial inhibition of hormone sensitive lipase in mice and in human adipocytes led to improved insulin resistance and glucose uptake (41).

### Adolescents with obesity may have specific microbiome characteristics based on 16S rRNA gene sequencing

We found significant differences in gut microbial community composition and presence or absence of specific taxa when comparing gut microbial 16S sequences between our HWC and OB interim cohort. While the majority of studies comparing gut microbes between OB and HWC have examined samples from adults, there are a few examples of studies that compare microbial consortia between obese and HWC children. A recent cross sectional study of 36 HWC and 42 Italian children with obesity aged 6-16 years (42) revealed that several individual microbes identified to their lowest taxonomic identifier correlated significantly either positively or negatively with an increasing BMI *z*-score (BMI normalized to age and sex), including *Bacteroides vulgatus* (R = −0.4321), *Faecalibacterium praustnitzii* (0.3508), and *Bacteroides stercoris* (−0.3252). A second study that examined differences between 81 HW, 29 overweight, and 80 OB children aged 9-11 years also found that *Faeacalibacterium* sp. was elevated with obesity, along with members of the Lachnospiraceae family (43), as we also demonstrated here. Further, our study includes a racially and ethnically diverse group of adolescents, which we hope will help to add important insights that may eludicate the factors contributing to a higher prevalence of pediatric obesity in nonwhite populations (44). A recent analysis of microbiome data from obese subjects found that race and ethnicity are major drivers of microbiome diversity in humans, and low representiation of samples from diverse subjects might help explain varied outcomes when describing the microbiome phenotype in obesity (12).

One goal of our work is to identify microbes associated with healthy weight in youth that could be developed as probiotics to augment current standard of care. An early clinical trial with a single microbe to treat obesity in adults has promising results in that supplementation over 3 months appears safe and effective in reducing markers of inflammation and liver disfunction (45). These data suggest that microbial-based therapies are promising additions to the arsenal of current obesity treatments in adults, and our data will help extend probiotic trials to adolescent patients who may not respond to standard care.

We still lack evidence to connect the adolescent obese microbiome with metabolic changes associated with youth onset of obesity and the related metabolome. The data reported here on the interim set of participants was not sufficiently powered to make these connections, however we will have the ability to analyze microbial contributions to the metabolome in the samples extracted from our full POMMS cohort of nearly 300 individuals. Overall, our preliminary data suggest that adolescents with obesity inhabit a transitional space in which some indicators of health appear normal, while others may be strongly predicative of later life disease development. Future analyses of microbiota in human and animal studies will help us define targets for new adolescent medicine therapeutics to intervene in what may otherwise be an unhealthy trajectory into adulthood.

## CONCLUSION AND IMPACT

Early life obesity increases the lifetime risk for cardiovascular disease and other obesity-related diseases. The proposed research will directly address a critical gap in obesity treatment for children: how to deliver the recommended ≥26 hours of treatment in a 6-month period to diverse groups in a way that is effective and sustainable. Our interim analysis suggests that microbiome and metabolome resesearch in adolescent obesity mirrors some results in adult obesity, but also offers new targets for further research, specifically surrounding BCAA-related metabolism and investigating the role that specific microbial taxa have in association with obesity or healthy weight. This clinical-basic science collaboration has resulted in a diverse sample set that can be used to better understand metabolomic-microbiome profiles in the development of obesity.

## Data Availability

All data referred to in the manuscript is either included as a supplemental file or has been uploaded to SRA/github. Some links are still in progress.

## Sources of Funding

National Institutes of Health R24-DK110492

American Heart Association 17SFRN33670990

## Acknowledgements

We would like to thank the study participants and their families for their involvement in this study. All raw data sequencing data will be made available via the NIH Sequence Read Archive (SRA) (submission in progress). Information on how to request access to the clinical sample biorepository and deidentified participant data can be found at https://sites.duke.edu/pomms.

